# Rest the Brain to Learn New Gait Patterns after Stroke

**DOI:** 10.1101/2024.04.01.24304938

**Authors:** Chandramouli Krishnan, Thomas E. Augenstein, Edward S. Claflin, Courtney R Hemsley, Edward P. Washabaugh, Rajiv Ranganathan

**Affiliations:** Department of Physical Medicine and Rehabilitation, Michigan Medicine; Department of Robotics, University of Michigan; Department of Mechanical Engineering, University of Michigan; School of Kinesiology, University of Michigan; Biomedical Engineering, University of Michigan; Department of Physical Therapy, University of Michigan-Flint; Department of Biomedical Engineering, Wayne State University; Department of Kinesiology, Michigan State University, East Lansing

**Keywords:** skill acquisition, error-based learning, motor task, hemiparesis, consolidation

## Abstract

**Background:** The ability to relearn a lost skill is critical to motor recovery after a stroke. Previous studies indicate that stroke typically affects the processes underlying motor control and execution but not the learning of those skills. However, these prior studies could have been confounded by the presence of significant motor impairments and/or have not focused on motor acuity tasks (i.e., tasks focusing on the quality of executed actions) that have direct functional relevance to rehabilitation.

**Methods:** Twenty-five participants (10 stroke; 15 controls) were recruited for this prospective, case-control study. Participants learned a novel foot-trajectory tracking task on two consecutive days while walking on a treadmill. On day 1, participants learned a new gait pattern by performing a task that necessitated greater hip and knee flexion during the swing phase of the gait. On day 2, participants repeated the task with their training leg to test retention. An average tracking error was computed to determine online and offline learning and was compared between stroke survivors and uninjured controls.

**Results:** Stroke survivors were able to improve their tracking performance on the first day (p=0.033); however, the amount of learning in stroke survivors was lower in comparison with the control group on both days (p≤0.05). Interestingly, the offline gains in motor learning were higher in stroke survivors when compared with uninjured controls (p=0.011).

**Conclusions:** The results suggest that even high-functioning stroke survivors may have difficulty acquiring new motor skills related to walking, which may be related to the underlying neural damage caused at the time of stroke. Furthermore, it is likely that stroke survivors may require longer training with adequate rest to acquire new motor skills, and rehabilitation programs should target motor skill learning to improve outcomes after stroke.

## Introduction

Stroke is a major cause of adult disability worldwide, affecting millions of people each year.^1^ Common motor impairments after stroke include weakness on one side of the body,^2^ difficulty coordinating movements,^3, 4^ and loss of balance.^5^ These impairments often result in disabilities that restrict the mobility and independence of stroke survivors in their daily activities, which in turn highlights the need for effective rehabilitation techniques that can improve walking ability. Current approaches to gait recovery after stroke often involve task-specific training with assistive devices and interactive technologies.^6, 7^ However, despite their effectiveness, these methods are no more beneficial than conventional rehabilitation in most clinical trials.^8, 9^ Therefore, there is a critical need for new therapies that can facilitate gait recovery after stroke.

A key to developing effective rehabilitation interventions after stroke is through the application of motor learning principles. Although the importance of incorporating motor learning principles into stroke rehabilitation programs has been repeatedly emphasized,^10, 11^ there still remains a large gap in our understanding of how learning and rehabilitation processes are interlinked in clinical populations.^12^ There is some evidence that acquiring new skills can activate neuroplastic mechanisms in the brain and that the process of learning a new motor skill shares similarities with relearning lost motor skills following a stroke.^10, 13^ Therefore, studying motor learning deficits after a stroke can provide a better understanding of the specific mechanisms of neurophysiological recovery, which could aid in the development of more effective interventions.

However, the effect of stroke on motor skill learning is difficult to estimate, as there is limited research on this topic and previous research has yielded conflicting results. For example, some studies suggest that stroke primarily affects the processes underlying motor control and execution, while leaving the learning of motor skills intact.^14-17^ However, a recent study revealed that the extent of motor learning deficits following a stroke is dependent on the severity of motor impairment.^18^ It is important to note that a major challenge in establishing evidence of learning deficits is that performance deficits can be misinterpreted as learning deficits.^19, 20^ This is supported by the observation that error-based learning capacity in stroke survivors is comparable to neurologically intact adults when motor execution deficits are controlled for during the experiment.^14, 17^ However, many of these prior studies, for good reasons, have focused on goal or action selection (i.e., where to move to or what movement can achieve the chosen goal) with less emphasis on motor acuity (i.e., the quality of the executed movements).^21^ More importantly, the experimental tasks are often restricted to a single degree of freedom (DOF) movement, thereby making it challenging to generalize these findings to complex multi-DOF movements (e.g., gait) and limiting their functional relevance to rehabilitation. As a result, it is currently unclear how stroke affects motor learning and whether learning deficits are present in individuals with minimal impairment when performing functional lower-extremity tasks such as walking.

Therefore, the purpose of this study was to evaluate the extent of motor learning deficits in chronic stroke survivors using a functional leg motor skill learning task. To minimize the impact of paresis/weakness on our findings, we specifically recruited stroke survivors with minimal impairment. To comprehensively understand the effect of stroke on motor learning, we examined both online (i.e., changes that occur during practice within the same day) and offline (i.e., changes that occur after practice during periods of no practice between days) learning. To address the issue of task relevance to day-to-day activities, the task required participants to learn a gait pattern that required 30% greater hip and knee flexion during the swing phase, which has been previously shown to be highly relevant in rehabilitation training for addressing stiff knee gait after stroke.^22^ We hypothesized that stroke survivors with mild motor impairments would exhibit significant deficits in both online and offline learning and retention of motor skills during walking when compared with uninjured controls.

## Methods

### Participants

A total of 25 adults (10 individuals with stroke and 15 uninjured controls, Table 1) participated in this study. This sample size provided us with a power β > 80% to detect statistical significance with an effect size of 𝜂𝜂^2^= 0.3 (derived from our prior studies) at a significance level of α = 0.05 (computed in G*power 3.1.9.7). Participants in the control group were also part of a different study that investigated the effects of aging on motor learning.^23^ All participants were right leg dominant based on their preferred leg to kick a ball ^24^. Stroke survivors were included in the study if they (1) had a radiologically (CT or MRI) confirmed ischemic or hemorrhagic stroke at least 6 months prior to the study, (2) had no significant cognitive deficits (Mini-Mental State Examination [MMSE] Score ≥ 22), (3) had no documented major sensory, motor, or proprioceptive deficits, (4) were able to walk independently with or without assistive devices, (5) had no history of uncontrolled diabetes or hypertension, and (6) had no major orthopaedic issues or range of motion deficits. Control participants were included in the study if they (1) had no significant cognitive deficits (MMSE ≥ 22), (2) had no significant orthopaedic or neurological issues, and (3) had no history major medical conditions, including uncontrolled diabetes or hypertension. We measured the stroke survivors’ lower extremity motor impairment with the lower-extremity Fugl-Meyer scale (LE-FM, 32.1 ± 2.2, one participant was not measured). All participants were tested at a single laboratory within the University of Michigan and signed a written informed consent prior to participation that was approved by the University of Michigan Human Subjects Institutional Review Board.

**Table 1:** Demographics of participants. Mean ± standard deviation for age, mass, height and MMSE score have been reported. The MMSE score can range from 0 to 30.

| **group** | **Sex** | **Age (year)** | **Mass (kg)** | **Height (m)** | **MMSE score** |
| --- | --- | --- | --- | --- | --- |
| **Stroke** | 5 females, 5 males | 58.3 ± 8.9 | 76.5 ± 20.5 | 1.7 ± 0.1 | 28.3 ± 2.2 |
| **Control** | 10 females, 5 males | 65.3 ± 2.9 | 72.0 ± 13.2 | 1.7 ± 0.1 | 29.3 ± 0.8 |

### Experimental Protocol

Participants learned a foot-trajectory tracking task on two consecutive days that were separated by about 24 hours (Figure 1(A)). Participants performed this task with their affected leg while walking on a motorized treadmill that was set to move at a constant speed of 0.89 m/s (2 mph) and wearing the same foot- and leg-wear (i.e., shorts or spandex). The foot-trajectory tracking task required participants to adjust their hip and knee angles during the swing phase of walking to match a target trajectory projected onto a computer monitor placed in front of them. On both days, the experiment consisted of four phases: baseline, pre-test (Pre), training (Tr), and post-test (Post) (Figure 1(B)). During baseline, participants walked normally on the treadmill for one minute. During the pre-test phase, the participants performed the foot-trajectory tracking task and their initial performance on the task was evaluated. The training phase consisted of repeated practice of the foot-trajectory tracking task. Participants completed eight blocks of practice, with each block lasting one minute and separated by one minute of rest. In the post-test phase, the participants again performed the foot-trajectory tracking task and changes in target-tracking performance were assessed. For stroke participants, the more-affected side was used as the training leg (3 left leg and 7 right leg), and for control participants, the training leg for each participant was determined randomly (7 left leg and 8 right leg). In the post-test phase, the participants’ final performance was evaluated by assessing their final target-tracking error.

**Figure 1:**
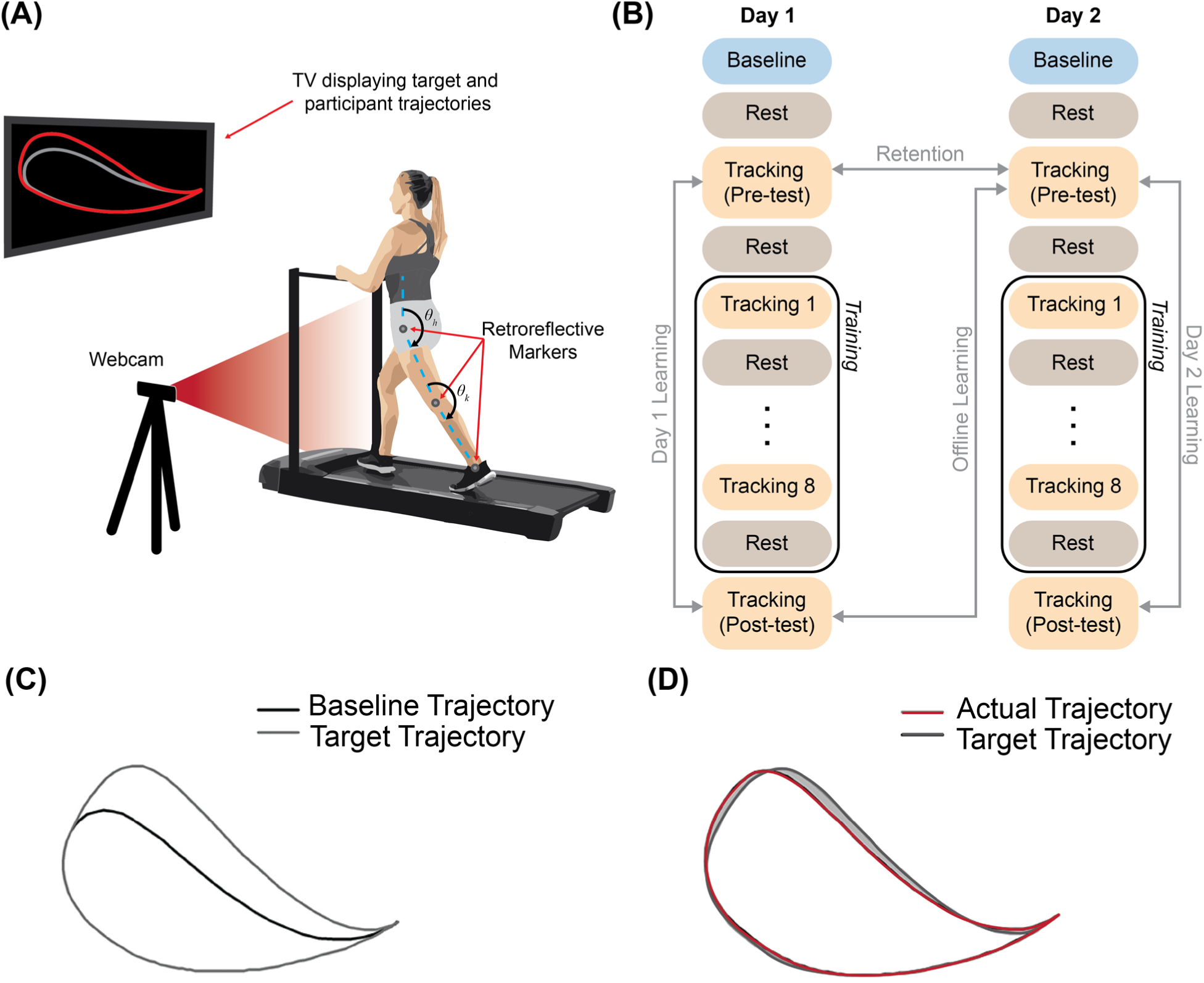
A schematic of the (A) experimental set-up and foot-trajectory tracking during treadmill walking, (B) experimental protocol, (C) participant’s baseline trajectory and their scaled (30%) target trajectory, and (D) computation of tracking error represented by the non-overlapping area (shaded in grey)

### Foot-Trajectory Tracking Task

A custom-designed, real-time motion tracking system, developed using LabVIEW 2011 and NI Vision Assistant (National Instruments Corp., Austin, TX, USA), was used for the motor learning task.^25^ The system consisted of a camera (C920 Pro HD Logitech Webcam, Logitech, San Jose, CA, USA) and computed the sagittal plane hip and knee kinematics during walking by tracking three 19 mm retroreflective markers positioned on the participants’ greater trochanter, lateral epicondyle of the femur, and lateral malleolus of the ankle. The target template trajectory for the foot-trajectory tracking task was created based on the participants’ sagittal plane hip and knee kinematics data obtained during baseline walking. The target trajectory was generated by scaling (1.3×) the hip and knee angles during swing phase of the normal walking trial and projecting this template in the end-point space, specifically the trajectory of the ankle relative to the hip on the sagittal plane (Figure 1(C)). This was achieved using the following forward kinematic equation:

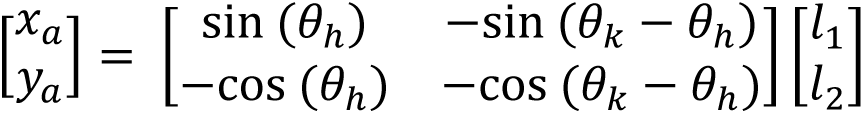

where 𝑥𝑥_𝑎𝑎_ and 𝑦𝑦_𝑎𝑎_ are the x and y positions of the ankle lateral malleolus relative to the hip, 𝑙𝑙_1_ is the distance between hip and knee markers (i.e., thigh segment), 𝑙𝑙_2_ is the distance between knee and ankle markers (i.e., shank segment), 𝜃𝜃_ℎ_ and 𝜃𝜃_𝑘𝑘_ are the anatomical hip and knee angles. The target template was smoothed using a Hanning window to prevent abrupt scaling at the beginning and end of the swing phase. The template trajectory was then displayed concurrently with the participant’s actual foot trajectory on a computer monitor positioned in front of the participant. Participants were instructed to try and match the target template trajectory as best as they can during the swing phase of their gait. Additionally, they were asked not to alter the normal gait patterns of the opposite leg that was not involved in the foot-trajectory tracking task.

### Data analyses

The performance on the foot-trajectory tracking task (i.e., how closely the participant’s actual trajectory matched the target trajectory spatially) was evaluated by computing the tracking error for each block. Tracking error was calculated as the difference in area (i.e., non-overlapping area) in pixels between the participant’s actual foot trajectory and the target template trajectory for each stride (Figure 1(D)). This stride-by-stride tracking error was then expressed as a percentage of the area within the participant’s target template and averaged across strides for each block. For the purposes of this study, four performance metrics were derived from the tracking error data on Day 1 and Day 2: (1) online learning (Day 1), (2) online learning (Day 2), (3) offline learning, and (4) retention. The amount of online learning on Day 1 (D1) and Day 2 (D2) was evaluated by comparing the tracking error during Pre blocks on Day 1 (D1-Pre) and Day 2 (D2-Pre) to Post blocks on Day 1 (D1-Post) and Day 2 (D2-Post), respectively. The amount of offline learning was evaluated by computing the difference in tracking error during the Pre block on Day 2 (D2-Pre) from the Post block on Day 1 (D1-Post). The amount of retention was evaluated by comparing the tracking error during the Pre block on Day 1 (D1-Pre) to Pre block on Day 2 (D2-Pre).

### Statistical analysis

All statistical analyses were performed in IBM SPSS for Windows Version 27 (SPSS Inc., Chicago, IL). A two-sample t-test was used to determine if there were any initial performance differences between the two groups. To evaluate if stroke affected the online learning and retention processes, we tested the differences in the amount of online learning (changes in tracking error on Day 1 and Day 2: D1-Post relative to D1-Pre and D2-Post relative to D2-Pre) and retention (initial tracking error on Day 2 relative to initial tracking error on Day 1: D2-Pre relative to D1-Pre) between the stroke and the control participants using repeated measures analysis of covariance (ANCOVA) with block as within-subject factor and group as between-subjects factor. To evaluate if stroke affected the consolidation process, we tested the differences in the amount of offline learning (changes in tracking error from the end of Day 1 to the beginning of Day 2: D2-Pre – D1-Post) between the stroke and the control participants using a one-way analysis of variance (ANOVA) with group as the between-subjects factor. These analyses are depicted graphically in Figure 1(B). A significant interaction effect was followed by appropriate post-hoc analysis with Sidak correction. Robustness checks were also performed by evaluating the results in relative terms (i.e., % baseline) in addition to the above method.^26^ For this analysis, we normalized the tracking error as a percentage of their respective baseline values (e.g., Day 1 online learning = [D1-Post/D1-Pre] × 100; offline learning = [D2-Pre/D1-Post] × 100) and compared those values between groups using two-sample t-tests with bootstrapping (10,000 iterations). A significance level of α = 0.05 was used for all statistical analyses.

## Results

### Day 1 Online Learning

Performance of a typical participant from each group is shown in Figure 2, and average group performance on the tracking task across each block on days one and two is shown in Figure 3. While there were no initial differences in tracking error between groups (t_1,23_ = 1.338; p = 0.194), there was a significant block × group interaction effect on the amount of online learning on Day 1 (F_1,22_ = 7.755; p = 0.011). Post-hoc analysis indicated that although both groups improved on tracking performance with practice on Day 1 (stroke: Δ = 3.2±1.4%, p = 0.033; control: Δ = 8.4±1.1%, p < 0.001; Figure 3B & Figure 4), the amount of tracking error at the end of practice on Day 1 was greater in stroke survivors when compared with the control group (19.6±1.4% vs. 14.5±1.1%, p = 0.033; Figure 3B & Figure 4).

**Figure 2:**
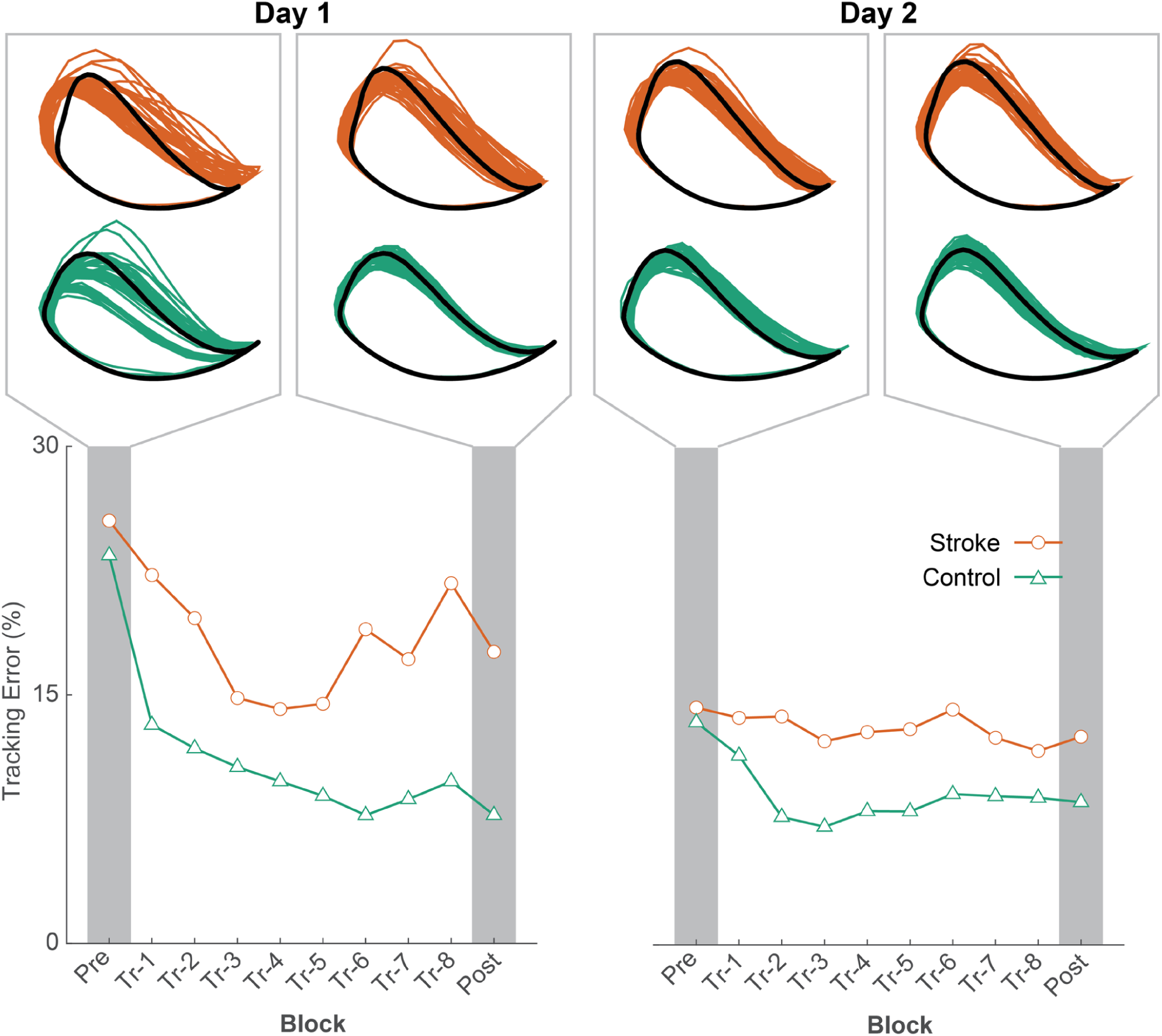
A representative example of participants’ tracking error in each group on Day 1 (left) and Day 2 (right)

**Figure 3:**
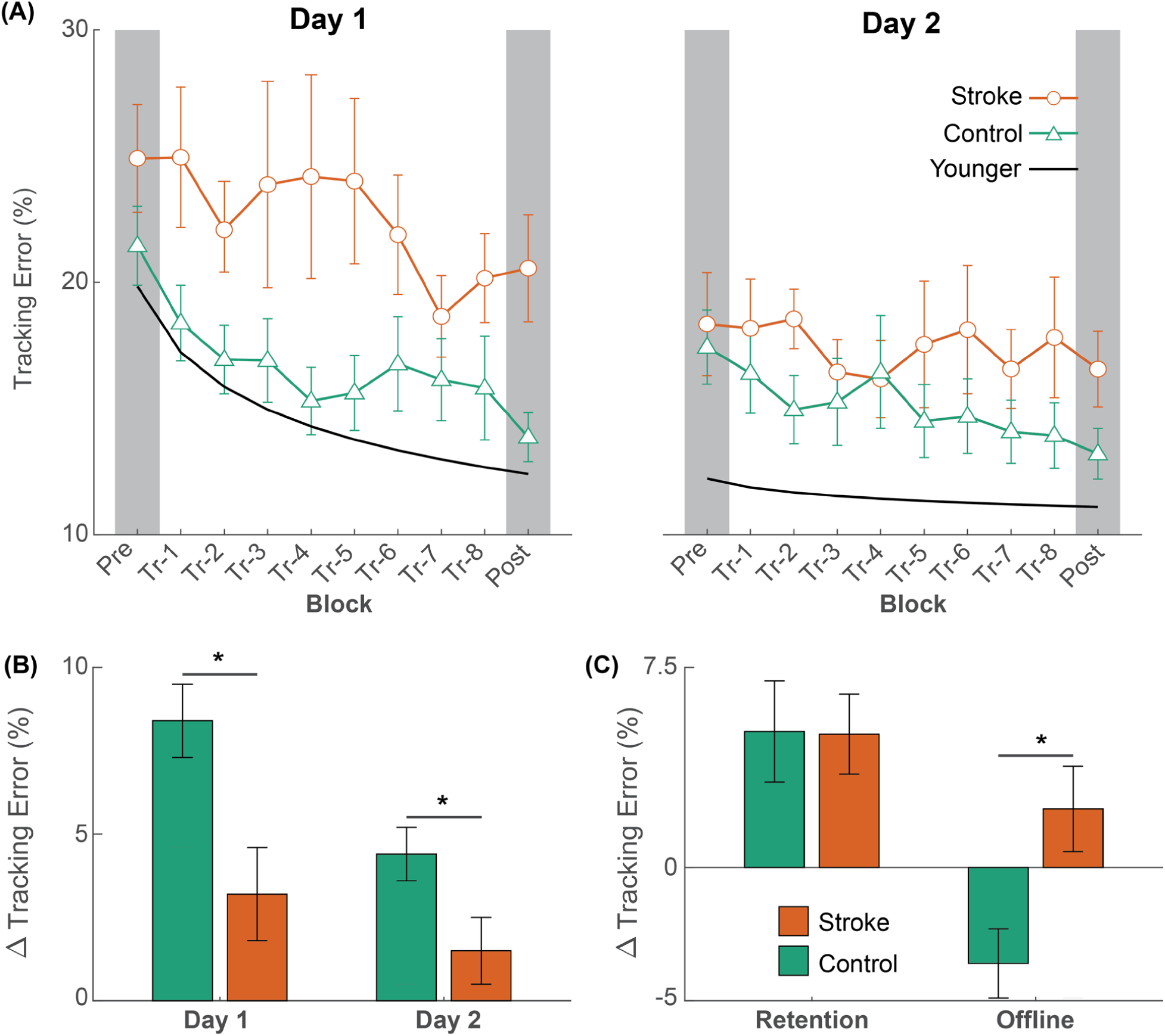
(A) The average trajectory tracking error in each group on Day 1 (left) and Day 2 (right). For comparison purposes, we provide data (power-fit curve of the mean data) from young, uninjured adults taken from a previous publication.^23^ **(B)** Bar plots showing online differences in learning between the stroke and the control group. **(C)** Bar plots showing differences in the amount of retention and offline gains between the stroke and the control group. Data for online learning and retention are shown as marginal mean changes (Δ) in tracking error. The error bars denote the standard error of the mean and asterisks (*) denotes statistical significance (*p* < 0.05). Positive values indicate improvements in performance.

**Figure 4:**
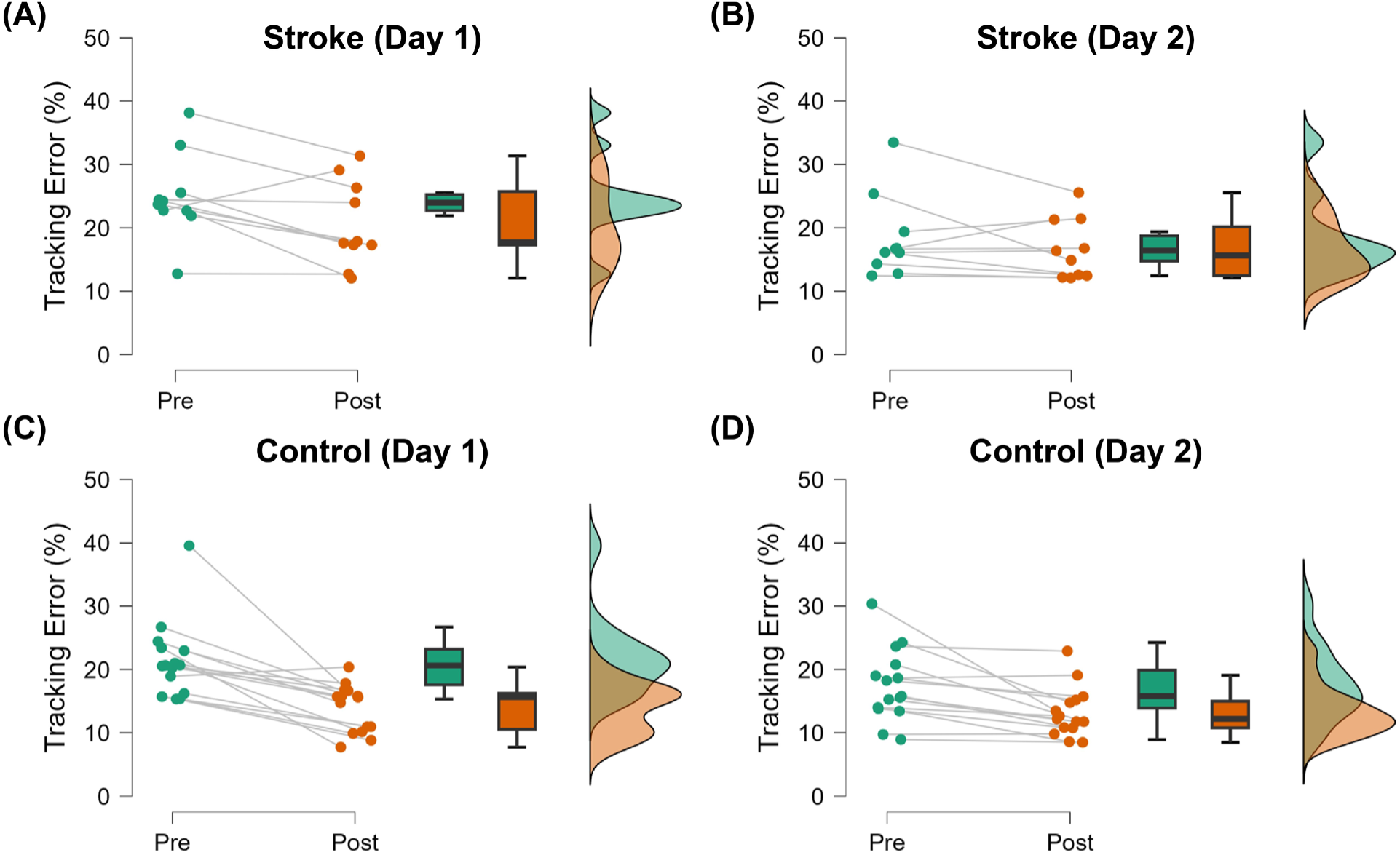
Raincloud plots showing distributions of normalized tracking error before (Pre) and after (Post) training in stroke survivors [top panel, **(A)** and **(B)**] and controls [bottom panel, **(C)** and **(D)**] on both days

### Day 2 Online Learning

There was a significant block × group interaction effect on the amount of online learning on Day 2 (F_1,22_ = 4.757; p = 0.040). Post-hoc analysis indicated that although the control participants improved on tracking performance with practice on Day 2 (Δ = 4.4±0.8%, p < 0.001), the stroke participants did not (Δ = 1.5±1.0%, p = 0.165). The amount of tracking error at the end of practice on Day 2 was greater in stroke survivors when compared with the control group (16.3±1.0% vs. 13.4±0.8%, p = 0.046).

### Retention

Retention of performance of the tracking task is shown in Figure 3C and Figure 5A. There was a significant effect of block on the amount of retention after training (F_1,22_ = 5.403; p = 0.030). On average, there was a 5.0±1.5% decrease in tracking error from D1-Pre to D2-Pre in stroke survivors and a 5.1±1.9% decrease in tracking error from D1-Pre to D2-Pre in the control participants. However, there was no group or block × group interaction effect on the amount of retention after training.

**Figure 5:**
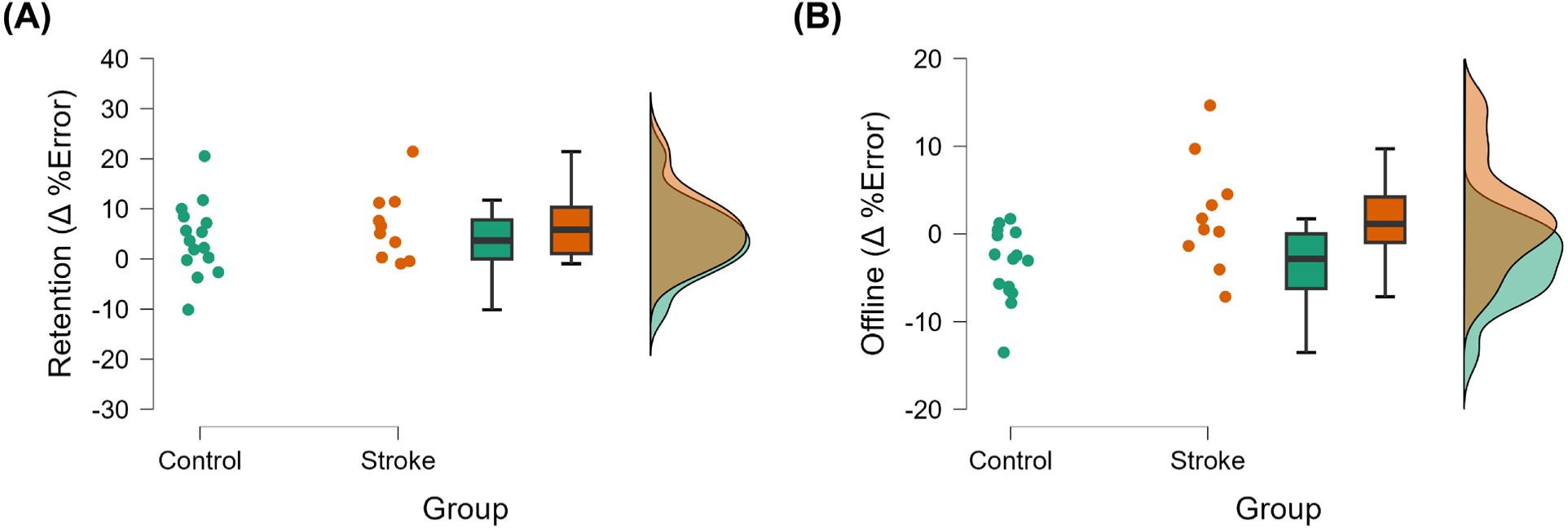
Raincloud plots showing distributions of **(A)** retention (computed as changes in normalized tracking error from Pre block on Day 1 to Pre block on Day 2) and **(B)** offline gains (computed as changes in normalized tracking error from Post block on Day 1 to Pre block on Day 2) in stroke survivors and controls

### Offline Gains

Offline changes in the performance of the tracking task are shown in Figure 3C and Figure 5B. There was a significant effect of group on the amount of offline gains in motor performance (F_1,23_ = 7.602; p = 0.011). On average, stroke survivors experienced a 2.2±1.6% reduction in tracking error from the end of Day 1 to the beginning of Day 2 (indicating offline gains), whereas control participants had a 3.6±1.3% increase in tracking error during the same period (indicating offline loss).

### Robustness Check

There was a significant difference between groups in the amount of online learning on Day 1 (stroke: 83.6±6.5%, control: 66.7±4.8%, mean difference = 16.9±7.5%, 95% bootstrapped confidence interval = 2.1–32.9%, p = 0.045), but the differences in the amount of online learning on Day 2 barely missed statistical significance (stroke: 93.2±6.1%, control: 78.6±4.4%, mean difference = 14.6±7.4%, 95% bootstrapped confidence interval = −0.1–29.1%, p = 0.066). There was also a significant difference between groups in the amount of offline learning (stroke: 92.7±8.0%, control: 127.7±8.1%, mean difference = −35.1±11.0%, 95% bootstrapped confidence interval = −56.9–−12.9%, p = 0.007) but no difference between groups in the amount of retention (stroke: 75.6±6.7%, control: 84.2±7.4%, mean difference = −8.6±9.6%, 95% bootstrapped confidence interval = −28.1–10.2%, p = 0.397). These results indicate that our findings were generally robust regardless of how the analyses were performed.

## Discussion

The objective of this study was to examine the extent of motor learning deficits in stroke survivors with low impairment using a functional, lower-extremity task that focused on motor acuity during gait. We focused on motor acuity because the bulk of literature examining motor learning following stroke has focused on the stroke survivor’s ability to select the right action (e.g., amplitude, order in sequence) rather than their quality of movement execution (e.g., kinematics, variability).^21^ Accordingly, stroke survivors and neurologically intact controls practiced walking on a treadmill with a new gait pattern in two separate sessions. To perform the new gait pattern, participants matched an ankle trajectory that necessitated 30% more hip and knee flexion during the swing phase. We found that stroke survivors showed a lower reduction in tracking error on both days (i.e., online learning deficits) when compared with the control group, who also are known to exhibit learning deficits due to the normal aging process (Figure 3).

Another key finding was that stroke survivors showed offline performance gains between days while the control group showed offline performance loss, indicating that although stroke survivors have lower ability in online learning, they have an advantage in offline learning during a period of rest. Although our study was limited by a small sample size, these findings were consistent regardless of how the analyses were performed (i.e., absolute vs. relative learning), indicating the robustness of our results.

The first notable finding from this study is that although stroke survivors were able to learn the task and improve with practice, they showed less improvement in their tracking error as compared with the control group on both days. This finding indicates that stroke survivors have online motor learning deficits when compared with neurologically intact adults, which agrees with some prior studies^27, 28^ but differs to some extent from other existing motor learning literature.^14-17, 29^ It is likely that this divergence from previous literature results from our examination of mildly impaired individuals using a multi-DOF, functionally relevant motor acuity task during gait. For example, recent research suggests that reinforcement learning was impaired early after the stroke but not in the chronic phase, whereas error-based learning was unaffected after stroke at either time point when compared with controls.^14^ However, our study indicates that online motor learning deficits are present even in chronic stroke survivors with minimal motor impairments when compared with controls. A key distinction between the two studies that could explain this discrepancy is the differences in the learning paradigm (skill learning in our study vs. visuomotor adaptation in the previous study). Further, gait is a highly practiced movement involving automatic processes such as balance and posture, and therefore invokes both conscious (*e.g.,* corticospinal) and automatic (*e.g.,* extrapyramidal) motor control pathways.^30^ For participants, learning a novel gait pattern requires making conscious alterations to this highly practiced movement, which could influence both these conscious and automatic pathways. On the contrary, non-functional upper-extremity tasks are generally unpracticed motions that do not impact balance and posture and are therefore more likely governed primarily by the conscious motor control pathways.^31^ Because these tasks likely require modulation of different motor control pathways, it is possible that they involve different learning mechanisms. Furthermore, even minimally impaired stroke survivors often have diminished balance that can influence their ability to prevent falls.^32^ Therefore, it is possible that the observed learning deficits arose from stroke survivors’ resistance to deviate from their current gait pattern and balance on one leg to perform the necessary exploration of the motor control task space, which is necessary to learn a motor skill.^33^

Another interesting finding was that despite the deficits in online learning observed in stroke survivors, we did not detect any differences in skill retention between the groups. This occurred because of the differences in offline learning between groups—stroke survivors exhibited offline performance gains whereas the control group exhibited an offline performance loss. This finding aligns with previous research highlighting the complex interplay between neurological damage and motor learning processes post-stroke.^10, 34^ One potential mechanism that could explain decreased ‘online’ learning is ‘reactive inhibition’ – i.e., stroke could increase reactive inhibition, which decreases online gains, but when the reactive inhibition dissipates, they “catch up” with the other group. This is typically observed in “massed” vs. “distributed” practice effects – the massed group shows poor online learning but then huge gains over the break.^35^

In our study, offline learning can be treated as a measure of sleep-dependent memory consolidation, i.e., a process of the central nervous system where recent memory traces are committed to long-term memory during rest.^36^ Existing research examining this process has shown that stroke survivors demonstrate greater motor performance in upper extremity motor tasks following a period of sleep as compared with an equivalent period of wakefulness, but the same is not true for neurologically intact individuals.^37-41^ Our results directly align with these prior findings and extend them to a functional lower-extremity task. This phenomenon may reflect heightened neural plasticity or alternative neural pathways recruited to compensate for damaged regions wherein post-stroke plasticity facilitates continued skill acquisition (or retention of skills acquired) during rest. Importantly, these findings indicate that stroke-induced neuroplastic changes can also lead to functionally beneficial adaptations apart from the commonly recognized maladaptive processes; albeit the precise mechanisms underlying these beneficial adaptations are not clear.

Several additional mechanisms could potentially explain why individuals with stroke may exhibit enhanced offline motor learning compared with older controls who do not demonstrate the same phenomenon. First, the brain undergoes significant neuroplastic changes after a stroke as it attempts to reorganize and compensate for the damaged areas.^20^ It is possible that these neuroplastic changes enhance the brain’s ability to consolidate and retain motor memories during rest or sleep, leading to improved offline motor learning.^34^ Second, stroke survivors often develop compensatory mechanisms (e.g., increased reliance on the premotor cortex or other undamaged regions) to overcome motor deficits.^42, 43^ During rest periods, these compensatory mechanisms may be reinforced or optimized, resulting in enhanced offline motor learning.^44^ Older controls, who have not experienced neurological damage, may lack the same need for compensatory mechanisms, and therefore, do not exhibit the same enhancement in offline motor learning. Third, stroke can disrupt normal sleep architecture, leading to alterations in sleep stages and patterns.^45, 46^ Some studies suggest that certain sleep stages, particularly REM and slow-wave sleep, are crucial for motor memory consolidation.^37, 47^ Changes in sleep architecture or enhanced sleep due to pharmacological effects (e.g., gabapentin improves slow wave sleep^48^ and total sleep time^49^) post-stroke may create a more conducive environment for offline motor learning compared with older controls. Finally, aging is associated with decreased neuroplasticity due to alterations in GABAergic activity,^50-52^ which may impact the brain’s ability to learn and consolidate motor memories during sleep. It is to be noted though that the differences in online and offline learning observed in this study occurred despite participants being in the chronic period of stroke recovery (i.e., when changes in neuroplasticity are believed to have plateaued), indicating that the enhancement in offline learning could be a sustained phenomenon. Overall, one or many of the above mechanisms likely contributed to the observed differences in offline motor learning between individuals with stroke and older controls. Further research is needed to fully elucidate the underlying mechanisms and their implications for stroke rehabilitation.

In summary, we investigated differences in learning a functional lower extremity motor skill between mildly impaired stroke survivors and neurologically intact individuals. We found that neurologically intact individuals showed greater motor performance with practice as compared to stroke survivors, but stroke survivors showed greater offline learning than neurologically intact individuals. These findings lend important insights into how stroke affects the learning process and may have potential implications for gait rehabilitation after stroke.

## Data Availability

Data will be available upon reasonable request to the corresponding author of the manuscript

## Acknowledgements

This work was partly supported by the National Institutes of Health (R41-HD111289) and the National Science Foundation Graduate Research Fellowship Program (DGE #1256260).

